# Cardiac MRI at 3.0T in the presence of implantable cardioverter defibrillators (ICDs)

**DOI:** 10.64898/2026.10.16.26364740

**Authors:** T. Reiter, O.M. Weber, I. Weiss, N. A. Kraus, M. Walther, S. Kress, W. R. Bauer

**Author notes:** Funding: Partial Funding was provided by SFB1525 grant number 453989101 and Biotronik SE & Co. KG, Berlin, Germany.

## Abstract

Patients with implantable cardioverter-defibrillators (ICDs) often are excluded from receiving CMR scans at 3.0T due to ICD-induced image artifacts. This work aims to transfer sequence protocols established for CMR imaging in the presence of ICDs at 1.5T to a first human application at 3.0T. All measurements were performed on a clinical 3T MRI scanner. Phantom measurements were used to establish and optimize wideband PSIR LGE imaging at 3.0T. Volunteer measurements with externally placed ICDs were used for optimization of CINE imaging and wideband PSIR imaging. Thirdly, patients with implanted ICD were examined with the optimized protocols. Human measurements showed that SA stacks obtained with spoiled GRE after contrast agent showed a markedly lower artifact burden than the conventional approach. For scar imaging, a wideband PSIR with a bandwidth of 3kHz and a frequency offset of +750Hz allowed a sufficient suppression of relevant artifacts in the majority of scans. No strong correlation between the image quality and the distance between the cardiac silhouette and the device was detected. The presented work shows the feasibility of clinical cardiac MRI at 3.0T including GRE-CINE and wideband PSIR-LGE for patients with ICDs.

## Introduction

Technical advances in active cardiac device therapy allow the safe examination of patients with MRI-conditional active cardiac implants within set conditions for the scanning environment and device setting. (1-4) However, the accessibility to MRI, especially CMR, remains limited for these patients. (5, 6) One aspect that accounts for this limitation are imaging artifacts that are induced by the implant and, to a lesser extent, the device leads. Generally, the induced artifacts have a variety of qualities such as changes in signal intensities, signal displacement and apparent changes in the size of structures. (7, 8) Due to the proximity of device to the region of interest, a significant overlap of artifacts across the myocardial structures can occur. (9) The type of active cardiac implants affects the overall artifact burden, with ICDs causing the largest burden. (10)

In clinical routine protocols, namely two sequences have been found to be problematic: the steady-state free precession sequence, routinely used for CINE imaging, is very susceptible to off-resonance effects. In the presence of a device, signal voids and signal pileup, along with geometric distortions and banding artefacts, frequently render the resulting images non-diagnostic. LGE imaging, on the other hand, conventionally employs an inversion pulse of typically limited bandwidth (1000 Hz). Frequency offsets in the presence of a device result in (regionally) un-inverted magnetization, potentially mimicking late enhancement and thus leading to false diagnosis.

The majority of current MR-conditional active cardiac implants allows scanning at a static magnetic field strength of 1.5T, but an increasing number of them also allow the use of 3.0T scanners. At 1.5T, changes in the imaging protocols such as usage of spoiled gradient echo sequences instead of SSFP have shown to be useful for CINE imaging. Additionally, late gadolinium enhancement (LGE) imaging in combination with a wideband inversion pulse has been established for artifact-free scar and fibrosis detection and so allows, for example, therapy assessment and planning for ventricular ablation procedures.(10-14)

Cardiac imaging at 3.0T has equally been established and is increasingly used for clinical studies. The higher field strength offers higher signal-to-noise ratios than at 1.5T. On the downside, the artifact burden increases as well, eroding the potential benefit of a higher field strength. This is especially apparent in the presence of medical implants. Literature on adaptation of imaging sequences on 3T, however, is scarce.

In this work, we use phantom and volunteer measurements in order to establish an adapted 3T protocol including spoiled gradient echo (GRE) for CINE imaging and wideband LGE-Phase Sensitive Inversion Recovery (PSIR). Contrast enhanced imaging sequences, both for CINE and LGE imaging, were established in patients with externally placed ICD as well as in patients with implanted ICD. This work shows the feasibility of such a protocol and quantifies the diagnostic improvement for scar tissue and cardiac function as compared to conventional sequences

## Methods

The study was carried out in three steps. Firstly, phantom measurements were used to establish wide band PSIR LGE imaging at 3.0T and to optimize the sequence setting for suppression of hyperintense artifacts. Secondly, volunteer measurements with externally placed ICDs were used for optimization of CINE imaging and wideband PSIR imaging sequences. Thirdly, patients with implanted ICD and a clinical indication for CMR were examined with the optimized protocols. All measurements were performed on a clinical 3T MRI scanner with commercial anterior (16 elements) and posterior (12 elements) body surface coils for signal reception (AchievaDS, dStream Whole body, Philips Healthcare, The Netherlands).

### Phantom measurements

For the phantom measurements, a previously described cubic phantom (28x28x28 cm) with a spherical grid system was used.(7, 8) Between the rows of spheres, small fluid-filled tubes with shorter T1 times were mounted.

Measurements were performed without and with a cardiac device (Ilesto 7 HF-T; Acticor 7 HF-T QP, Biotronik GmbH Co. KG) placed at the center of the phantoms. Adiabatic inversion pulses with bandwidths of 1 kHz, 2 kHz, 3 kHz and 4 kHz, and frequency offsets from -1000 Hz to +1500 Hz were tested. Inversion time was set to 110 ms based on a previously performed Look Locker sequence. Images were analyzed regarding artifact distribution and adequate signal suppression of the fluid-filled tubes.

### Human measurements

For in vivo evaluation of the tested protocol settings three groups of volunteers and patients were included (Tab 1). In short, the first group included healthy volunteers, the second, volunteering patients without implanted ICD who were scheduled for a clinical CMR scan, and the third patients with implanted ICD and indication for a clinical CMR scan. Approval from the local ethics committee was obtained for all human measurements, and all measurements adhered to relevant guidelines and regulations. Written informed consent was obtained from all participants. Monitoring during the CMR scan was performed according to current recommendations and included, for all patients with implanted ICD, continuous pulsoxymetry and repeated blood pressure measurements. (4) Vector ECG proved to be highly distorted in the presence of an ICD and did not serve as reliable monitoring tool. Device programming was performed according to vendor and guideline recommendations immediately prior and after the CMR scan. When the ICD was deactivated, trained medical personnel was always present.

**Tab 1:** Study population and tested sequences.

| Group | Number<br>(Male/female) | Mean age<br>(range) | Mean weight<br>(range) | Contrast agent<br>(Gadobutrol,<br>0,1 mmol/kg) | Clinical indication<br>for CMR | Device position | Protocol overview |
| --- | --- | --- | --- | --- | --- | --- | --- |
| Group I | 8 (4/4) | 26 years<br>(20-30 years) | 68 kg<br>(55-95 kg) | No | No<br>healthy volunteers | Externally taped,<br>left upper<br>pectoral region | CINE imaging (SA, 4CH-, 2CH-view);<br>bSSFP and GRE<br>flow compensation |
| Group II | 9 (8/1) | 67 years<br>(42-83 years) | 88 kg<br>(67- 111 kg) | Yes | Yes<br>patients | Externally taped,<br>left upper<br>pectoral region | Clinical protocol (CINE, PSIR LGE)<br>Study protocol (20- 25 min after contrast<br>agent)<br>Wideband PSIR LGE<br>CINE GRE (SA, 4CH-, 2CH-view) |
| Group III | 16 (13/3) | 64,5 years<br>(46–85 years) | 90 kg<br>(50-112 kg) | Yes | Yes<br>patients | Implanted,<br>left sided (14/16)<br>or<br>right sided (2/16) | Pre and post contrast agent:<br>CINE GRE (SA, 4CH-, 2CH-view)<br>Wideband PSIR LGE (after 10- 15 min) |

The tested sequences for each group are listed in Tab. 1. In short, in the group with healthy volunteers, native CINE imaging with different sequence types was tested. In both patient groups the focus was on late gadolinium enhancement and contrast enhanced CINE imaging. In group II, both wideband PSIR and CINE GRE imaging were obtained after completion of all necessary clinical scans. In group III, CINE imaging was obtained during the waiting period between administration of contrast agent and wideband PISR imaging. Sequence parameters are given in Tab 2. Wideband PSIR LGE with an adiabatic inversion pulse and variable bandwidths were used for both phantom and human measurements. In short, an adiabatic inversion pulses invert magnetization reliably even in the presence of B1 variations across the imaging volume, making them less sensitive to field inhomogeneities and leading to improved inversion uniformity, at the cost of higher RF power and SAR, longer RF pulses, and higher hardware demand. (Hwang TL Fast broadband inversion by adiabatic pulses. J Magn Reson. 1998 Jul;133(1):200-3)

**Tab 2:** Sequence settings.

| CINE Imaging |  |  |
| --- | --- | --- |
| - bSSFP | TR: 2,62 ms; flip angle 30°; Number of phase encoding steps: 218; Echo train length: 17; Pixel bandwidth: 2484; partial fourier direction: in phase; number of frequency encoding steps 176; no parallel acquisition; FOV 300 – 350 mm, resolution 1.8 – 2.0 mm, reconstruction resolution 1 mm, slice thickness 8mm. SAR 2,178-2,381 W/kg; no flow compensation. |  |
| - spoiled GRE (pre/post contrast) | TR: 2,38-5,02 ms; flip angle: 10°-19°; Number of phase encoding steps 166-208; Gradient echo train length: 10-18; Pixel bandwidth 542-1127; partial fourier direction: in phase or combination; number of frequency encoding steps 176; FOV 300 – 350 mm, resolution 1.8 – 2.0 mm, reconstruction resolution 1 mm, slice thickness 8mm; SAR 0,41-2,02 W/kg; with and without flow compensation. |  |
| Late gadolinium enhancement |  |  |
| (Wideband) PSIR LGE | TR 4,02 ms; TE 1,93 ms; Flip angle 15°, Inversion Time 365 ms; Number of phase encoding steps 175; Echo train length 41; Pixel bandwidth 434; number of frequency encoding steps 196; FOV 300 – 350 mm, resolution 1.8 – 2.0 mm, reconstruction resolution 1 mm, slice thickness 8mm, SAR 0,48 W/kg.<br>Bandwidth 1 up to 4 kHz, frequency offset -1500 to +1500 Hz | In group II 20- 25 min after contrast agent<br>In group III 10- 15 min after contrast agent |

In group II (patients without implanted device), the clinically indicated protocol was completed prior to placing an ICD externally on the chest. The number of sequences tested in each patient was limited according to the overall duration of the examination and the patient’s tolerance of the taped ICD.

For study group 3 (patients with implanted ICD), the overall imaging protocol was limited to 30 to 35 min. Survey scans in axial, coronal and sagittal orientation were used for planning the standard views (SA, 4- and 2-chamber views as needed). For CINE imaging the previously established GRE sequences were used prior and post administration of contrast agent. A Look Locker sequence was used for defining the optimal delay time, and wideband PSIR LGE imaging for detection of scar or fibrosis. Seven patients received conventional and wideband-PSIR LGE imaging with full ventricular coverage for both sequence types. In the remaining eight patients, the imaging protocol did not include conventional PSIR imaging.

### Image interpretation

The distance between the heart and the implanted device was measured on p.a. thoracic x-ray images using the unobstructed presentation of device and cardiac silhouette. The shortest distance between the device edge and the cardiac silhouette as well as the distance between the tip of the ventricular lead and the device were measured. (15) Measurements on the MRI Survey scans were omitted due to the severe artifact burden and lack of reproducible measurement endpoints. (15-18)

CMR image interpretation was performed by two CMR experts (TR, WB, both > 10 years CMR experience). The analysis and interpretation of all clinically indicated scans (group II and III) followed the 2020 SCMR recommendations. (19) All full short axis stacks (full ventricular coverage) were segmented manually. The endomyocardial border was traced in end-diastole and end-systole, and LV ejection fraction calculated according to the Simpson’s method. Artifact burden was assessed qualitatively in both SSFP and GRE post contrast CINE images regarding presence of ripple artifacts and signal void overlapping with myocardial structures. All slices of each full short axis stacks were analyzed regarding presence and quality of artifacts. A Pearson correlation coefficient with the shortest measured distance from the device to the cardiac silhouette was calculated.

Similarly, the late gadolinium enhanced images were qualitatively analyzed and all affected slices in conventional and wideband PSIR LGE short axis stacks counted. Additionally, in full axis scans, the visible hyperintense area was traced manually and the overall artifact volume was calculated from all slices. Hyperintense artifacts that did not affect myocardial structures at all were not included. Pearson correlation coefficient with the measured distance was calculated as well.

## Results

### Results CINE Imaging

The CINE measurements in the volunteer group demonstrated the expected high artifact burden of CINE imaging with bSSPF sequences, regardless of different sequence settings. A spoiled GRE sequence (TR 3 ms, TE 1 ms, flip angle 15°, echo train length 49 with flow compensation and partial echo) showed the best suppression of ripple artifacts. However, the blood pool remained inhomogeneous and the endomyocardial border appeared slightly blurred. After application of contrast agent, these effects were reduced (See supplementary figure 1).

These sequence settings were maintained for measurements in the group of patients with implanted device (group 3). In 8 patients with left sided implant, short axis scans were performed using the conventional bSSFP and post-contrast spoiled GRE sequences. 78% to 100% of all slices obtained with bSSFP showed a significant ripple artifact burden, with a focus on the anterior, septal and lateral segments. Opposed to these findings, SA stacks obtained with the spoiled GRE after contrast agent showed a markedly lower artifact burden (Fig. 1).

**Fig 1:**
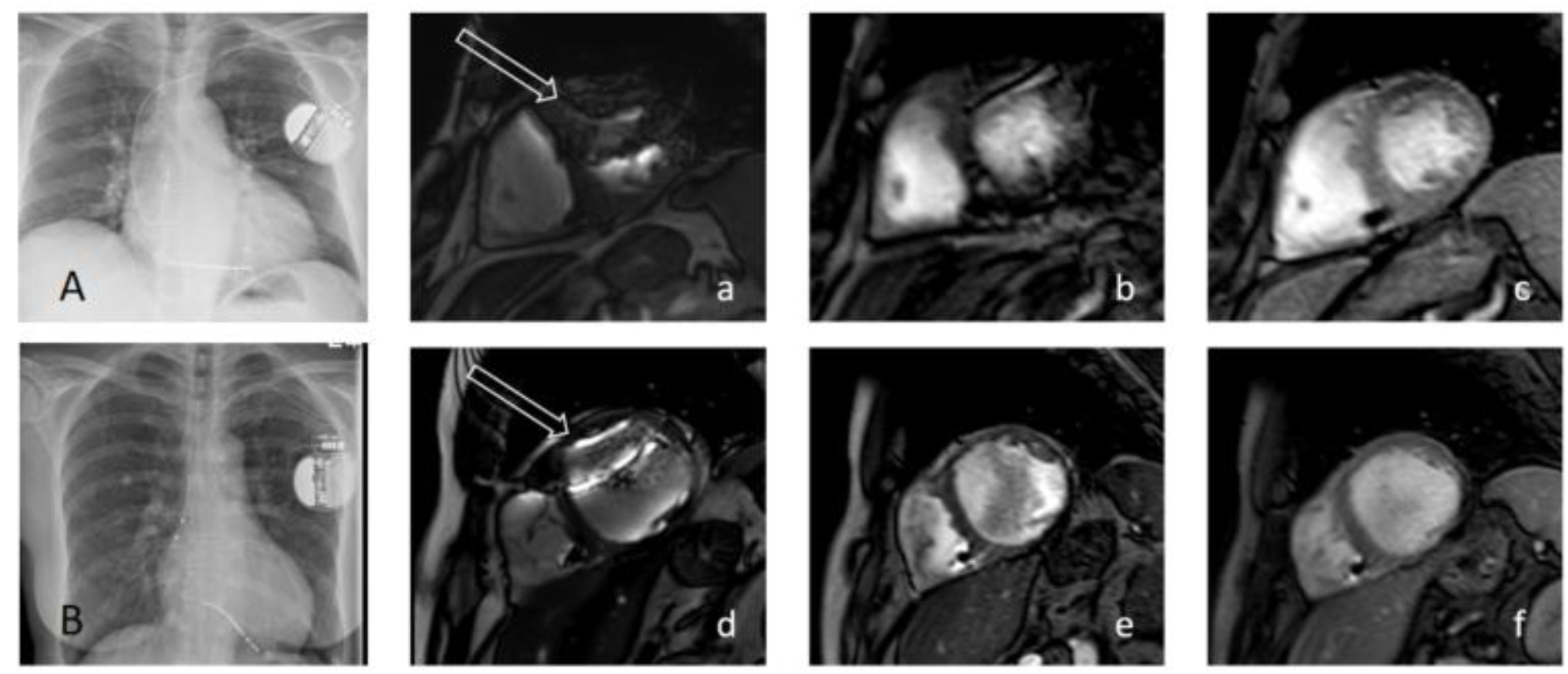
Comparison of bSSFP, native spoiled GRE and contrast enhanced spoiled GRE CINE imaging in two patients with implanted ICD. Patient A (a-c) with DCM: A) shows the p.a. x-ray image, distance device-cardiac silhouette 30 mm a)extensive ripple artifacts caused by bSSFP imaging overlap with septal, anterior and lateral segments (white arrow). b) native spoiled GRE suppresses the majority of ripple artifacts, however, the endomyocardial border appears blurred. c) artefact free short axis view obtained with contrast enhanced spoiled GRE imaging. Patient B (d-f) with suspected sarcoidosis. B) B) shows the p.a. x-ray image, distance device-cardiac silhouette 42 mm d) bSSFP imaging causes ripple artifacts (white arrow), however, the majority of segments are clearly visible. e) native spoiled GRE imaging suppresses these ripple artifacts and the myocardium can be clearly distinguished from the blood pool. f) contrast enhanced spoiled GRE imaging allows a better presentation of the endocardial trabeculae.

CINE imaging with contrast enhanced spoiled GRE imaging avoided ripple artifacts regardless of the distance between the device and the cardiac silhouette (r^2^= 0.001) (Fig. 2). In three cases, an overlap with a signal void was present, and in one of these cases, the device was located in a particularly short distance (38 mm) from the cardiac silhouette. In this case, the signal void overlapped with the myocardium rendered only 34% of slices without relevant artifacts (See Supplementary Figure 2). Notably, the artifact burden in the bSSFP scans also showed no correlation with the distance between device and cardiac silhouette (r^2^= 0,04), however, with this sequence type, the vast majority of slices showed relevant artifacts.

**Fig 2:**
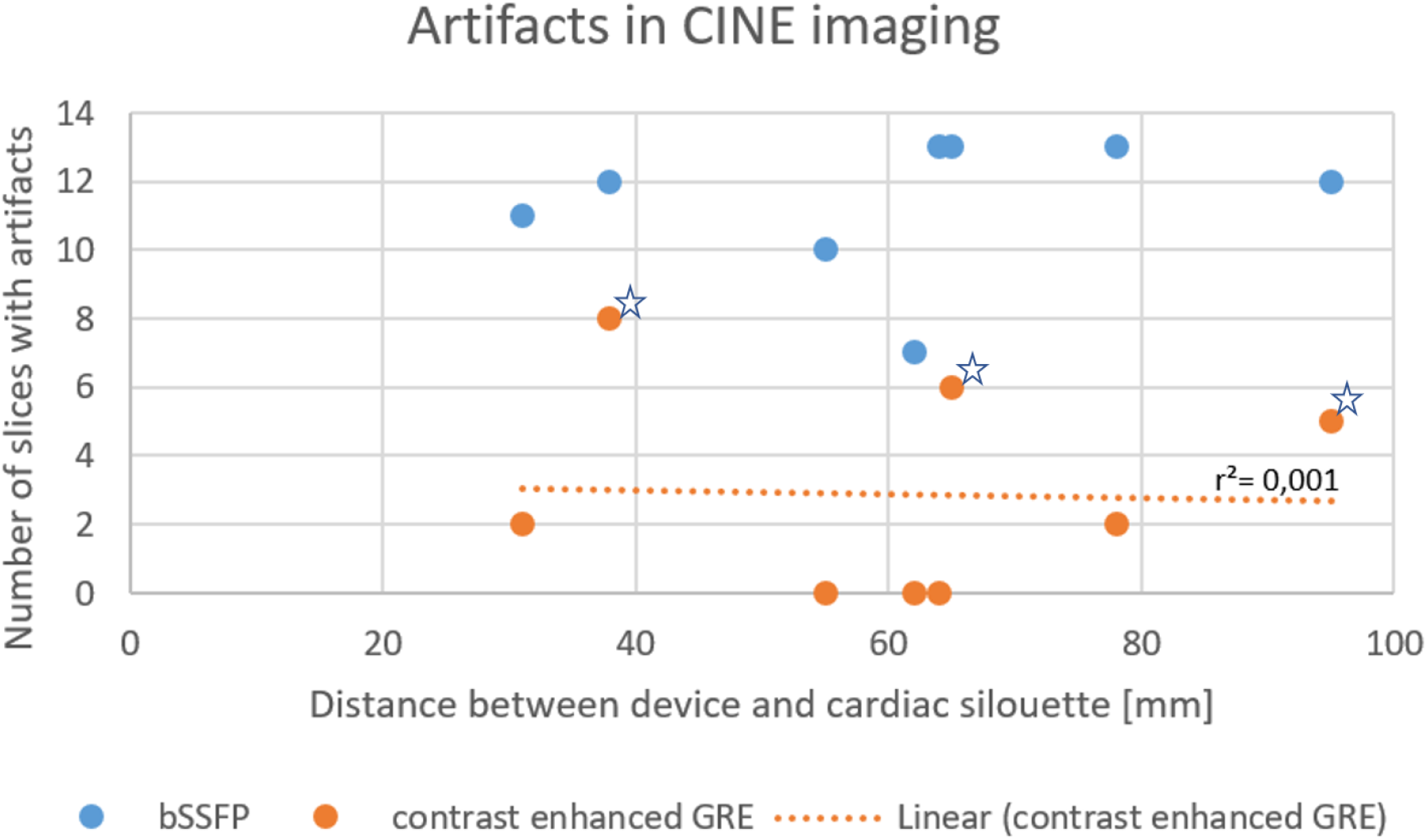
Comparison of bSSFP and contrast enhanced GRE. The artifact in the marked three contrast enhanced GRE sequences were signal voids (Asterisk). In these three scans, no ripple artifacts were present.

### LGE measurements

#### Phantom measurements

In the presence of an active implant, a variety of artifacts such as hyper- and hypo-enhancement as well as displacement artifacts were visible. The efficacy of the wideband inversion pulse for suppressing these artifacts varied with bandwidth and frequency offset. A frequency offset of 0 Hz had little effect on displacement artifacts, whereas a positive frequency offset reduced the artifacts. In the phantom setting, the comparison of different bandwidths and frequency offsets showed that the best results were obtained with a bandwidth of 3 kHz and an offset of +750 Hz (see supplementary figure 3).

#### Human measurements

All patients of group 2 completed the additional measurements with the externally placed ICD without complication. The device-induced artifacts were detectable in all subjects. However, due to the external placement of the ICD, not all artifacts were seen in close proximity of the cardiac region. The pulse setting that had shown the best results during the phantom measurements, a bandwidth of 3kHz and a frequency offset of + 750 Hz, were tested and allowed a good suppression of hyperintense artifacts overlapping with the cardiac region. Existing lesions were adequately detected (Fig. 3).

**Fig 3:**
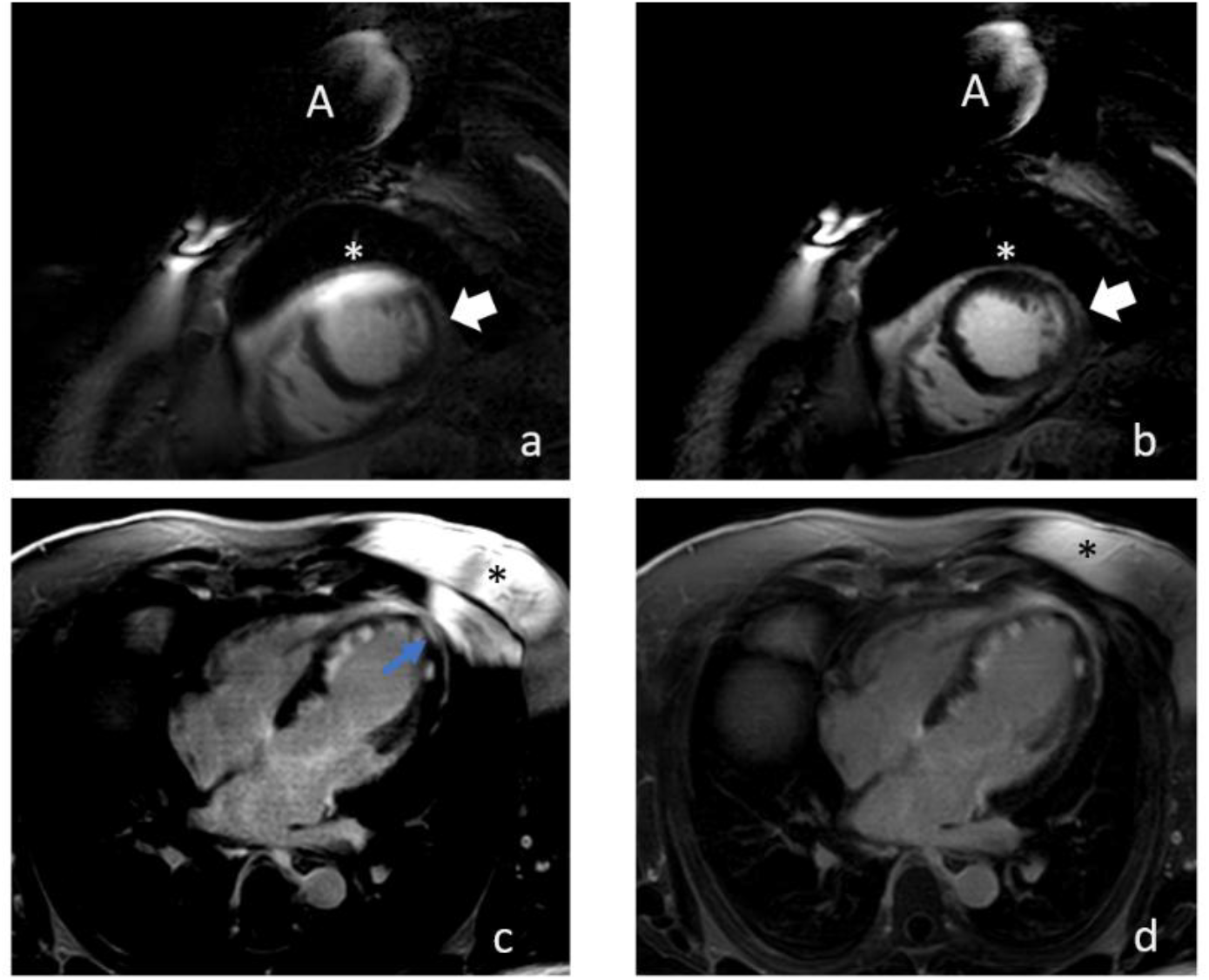
Patient measurements with an externally placed ICD. (A) marks the implant, visualized by the central signal void and hyperintense artifacts in the immediate proximity. a) and b) show short axis scans with conventional and wideband PSIR LGE. The asterisk (*) marks the hyperintense artifact overlapping the myocardium, the white arrow marks the actual existing subendocardial lesion. c) and d) show a four-chamber view with extensive subendocardial scarring after myocardial infarction. The asterisk (*) marks the hyperintense imaging artifact. The small region of overlap mimics an epicardial scar (blue arrow), that is not present in the measurement with the wideband pulse.

In the group of patients with implanted device (group III) one patient did not complete the CMR scan due to severe arrhythmia and contrast agent paravasate. This patient was not included in the further analysis. 60% (9/15) of the patients had an ICD as primary prevention, and (33%) 5/15 for secondary prevention; details are given in Tab 3. 40% (6/15) patients did not require any pacemaker activity, 40% (6/15) showed 1-5% RV pacing, and 20% (3/15) had 74% or more RV pacing.

**Tab 3:** Patient and device characteristics. Due to data privacy, age is given in ranges.

| Patient No. | Sex | BMI (kg/m<sup>2</sup>) | Vendor | Device | Leads | Primary/secondary Prevention | Leading Diagnosis | Indication CMR |
| --- | --- | --- | --- | --- | --- | --- | --- | --- |
| 1 | M | 31,6 | Biotronik | Rivacor 7 VR-T Dx DF4 ProMRI | Plexa S DX65; 17ProMRID F-1 | 1 | DCM | Substrate analysis |
| 2 | F | 18,1 | Medtronic | Visia VR MRI | Sprint Quattro MRI | 2 | Acute sarcoidosis | Inflammation |
| 3 | F | 33,4 | Biotronik | Intica 7VR-T DF4 Pro MRI | Plexa S65 ProMRI DF-4 | 1 | iCMP | Substrate analysis |
| 4 | M | 32,7 | Boston | Charisma VR 4 DF4 | Reliance 4 FR 64 cm | 1 | Suspected HCM | Substrate analysis |
| 5 | M | 28,1 | Boston | Charisma | Endotak Reliance | 2 | DCM | Substrate analysis |
| 6 | F | 20,8 | Biotronik | Rivacor 5 VTR-Dx | Plexa ProMRI S DSX 65/15 | 1 | Acute Sarcoidosis | Inflammation |
| 7 | M | 27,4 | Biotronik | Rivacor 5 VRT-Dx | Plexa ProMRI S DX 65/15 | 1 | VTs | Substrate analysis |
| 8 | M | 33,4 | Biotronik | Intica 7 Vr-T | Plexa ProMRI S65 | 1 | iCMP | Substrate analysis |
| 9 | M | 21,9 | Biotronik | Rivacor 5 VR-T DF4 | Plexa ProMRI S65 | 1 | DCM | Substrate analysis |
| 10 | M | 29,4 | Biotronik | Iforia7 VRT | Protigo ProMRI S65 | 1 | iCMP | Substrate analysis |
| 11 | M | 23,8 | Biotronik | Rivacor 5 DR-T DF4 proMRI | Plexa ProMRI S65 DF4 | 2 | DCM | Substrate analysis |
| 13 | M | 29,7 | Boston | Charisma EL | Ingevity MIR 52 cm; Reliance 4-front 64 cm | 2 | iCMP | Substrate analysis |
| 14 | M | 26,9 | Boston | Charisma EL | Endotak Reliance S | 2 | iCMP | Substrate analysis |
| 15 | M | 30,9 | Biotronik | Rivacor 5 HF-T | Solia S53; Protego ProMRI S53 | 1 | iCMP | Substrate analysis |
| 16 | M | 33 | Biotronik | Rivacor 7 VR-T | Plexa S65 ProMRI | 2 | iCMP | Substrate analysis |

The distance between the RV/Shock coil and all left sided devices ranged from 9.3 cm to 17.4 cm (median, 13.2 cm) and between the heart silhouette and left sided devices from 2.1 cm to 9.4 cm (median, 5.5 cm). The distance between the RV/Shock coil and the right sided devices was up to 16.7 cm and between the cardiac silhouette and the device 23.4 cm. LV function ranged from 12 to 58% (median, 40%), with an EDV from 124 to 386 ml (median, 218 ml) and a SV from 46 to 139 ml (median, 73 ml).

For LGE imaging, the initial pulse setting was a wideband pulse of 3kHz and an offset of + 750 Hz. The wideband pulse for PSIR LGE did not fully suppress the hyperintense artifact in three patients (See supplementary figure 4). In one of these patients, a probatory wider bandwidth (5 kHz) did reduce the artifact burden but did not fully suppress the hyperintense artifact. The right sided implants did not negatively impact image quality. Changes in the frequency offset resulted in shift of the artifact burden from the cardiac region (Fig. 4).

**Fig 4:**
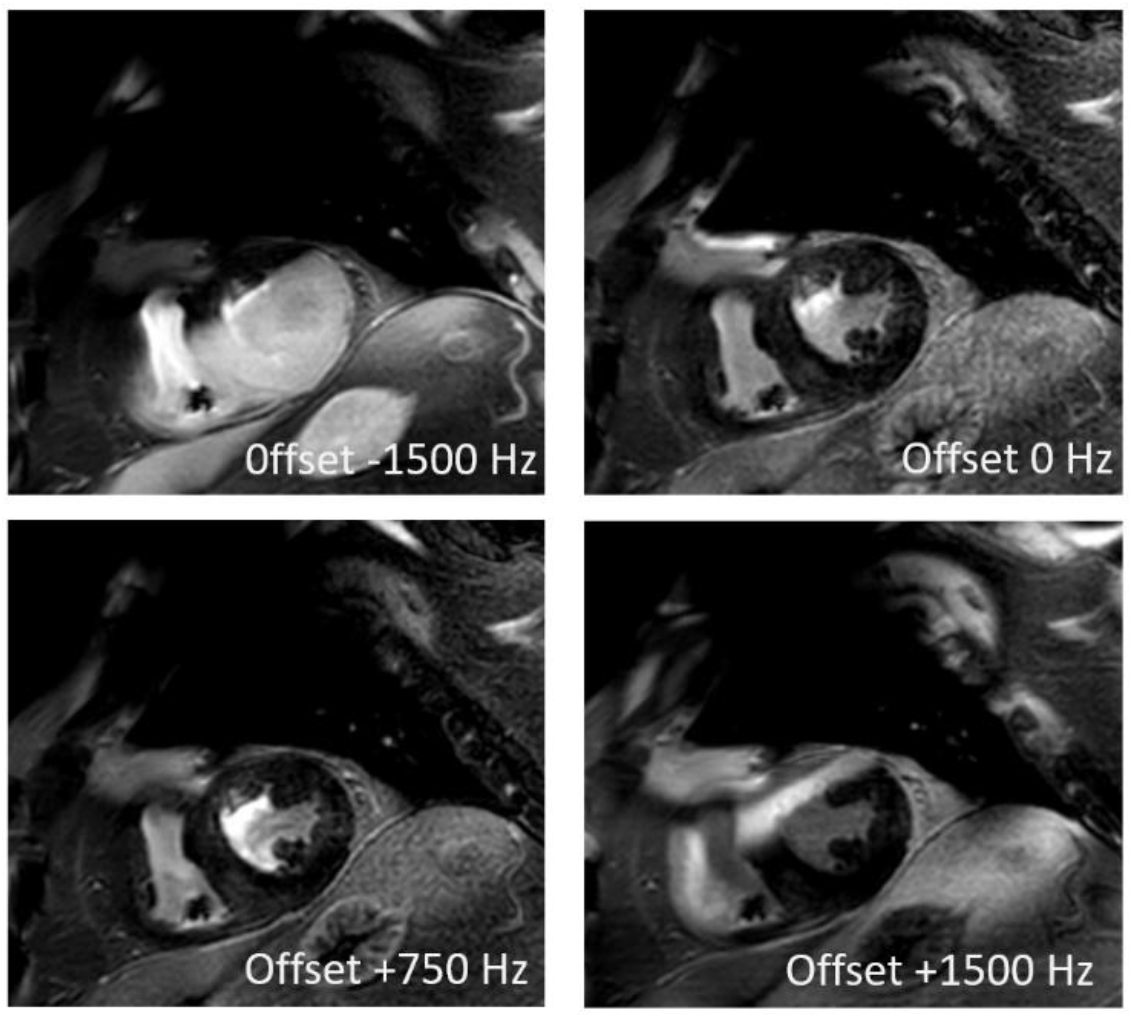
Wideband LGE PSIR imaging with a bandwidth of 3kHz. Effect of different frequency offsets on artifact distribution in a patient with implanted 1 chamber ICD. The localization the of hyperintense artifact switches with positive or negative offset.

In the sub-group of seven patients that were scanned both with conventional PSIR-LGE imaging and wideband PSIR-LGE imaging (bandwidth 3kHz, frequency offset + 750 Hz), the artifact burden was markedly lower when the latter sequence was used. In 5/7(71%) patients, the cardiac region was artifact-free, whereas with conventional PSIR-LGE only one of these seven patients (14%) had an artifact-free scan (Fig. 5). When present, the artifact volume was markedly lower for the wideband PSIR Imaging compared to the conventional PSIR imaging. In our series, a weak correlation with the distance between the device and the cardiac silhouette was detected (r^2^0= 0.31) (Fig. 6).

**Fig 5:**
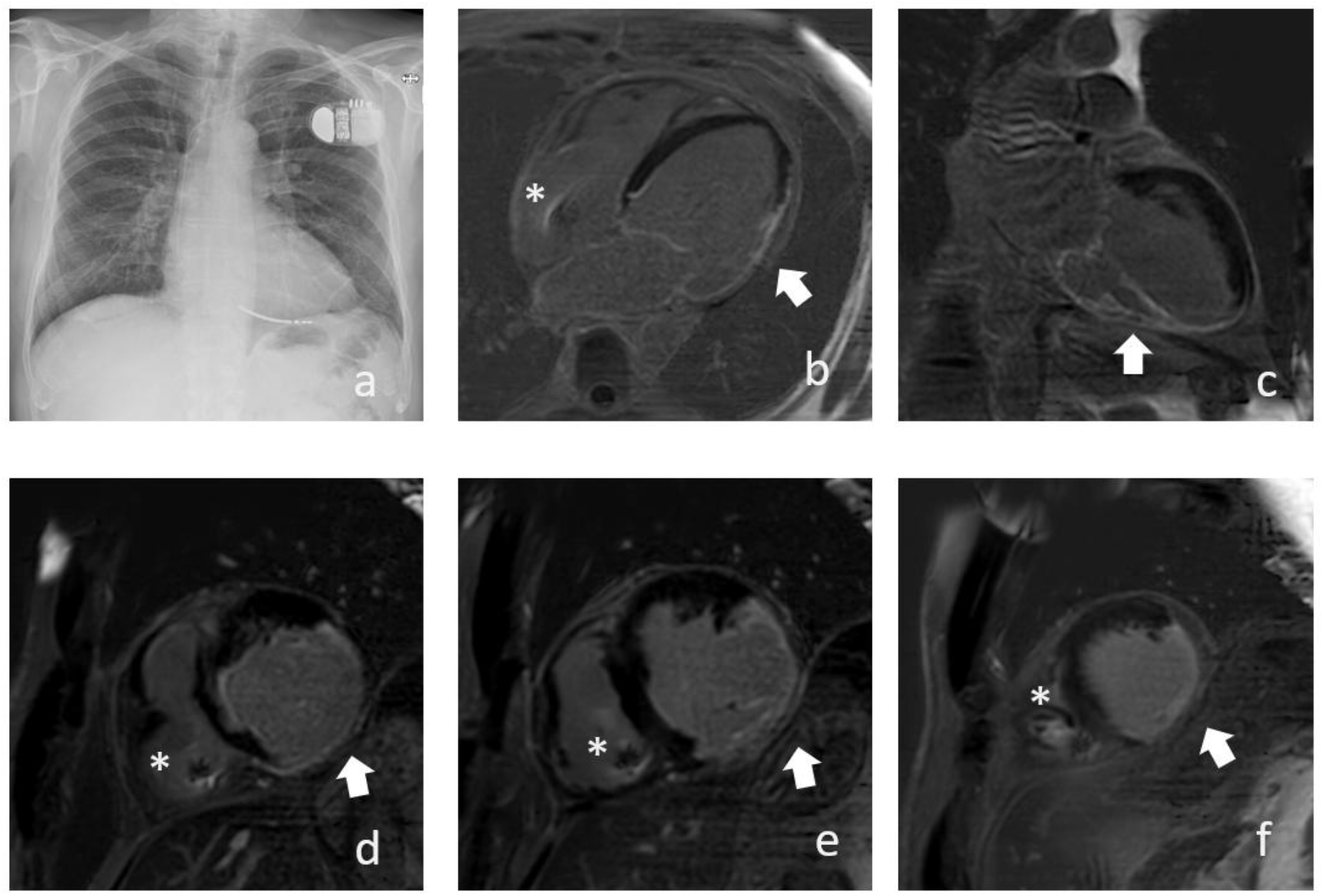
Patient with ischemic cardiomyopathy as result of a posterior wall infarction and implanted one-chamber ICD (distance device-cardiac silhouette 78 mm). Substrate imaging with Wideband PSIR LGE (Bandwidth 3kHz, Frequency offset +750Hz). a) Chest X-ray (pa). b) 4-chamber view c) 2-chamber view and d-f) short axis views (d) basal, e) midventricular, f) apical) with ischemic scar tissue posteriorly and laterally (arrow). Lead artifact (Asterisk)

**Fig 6:**
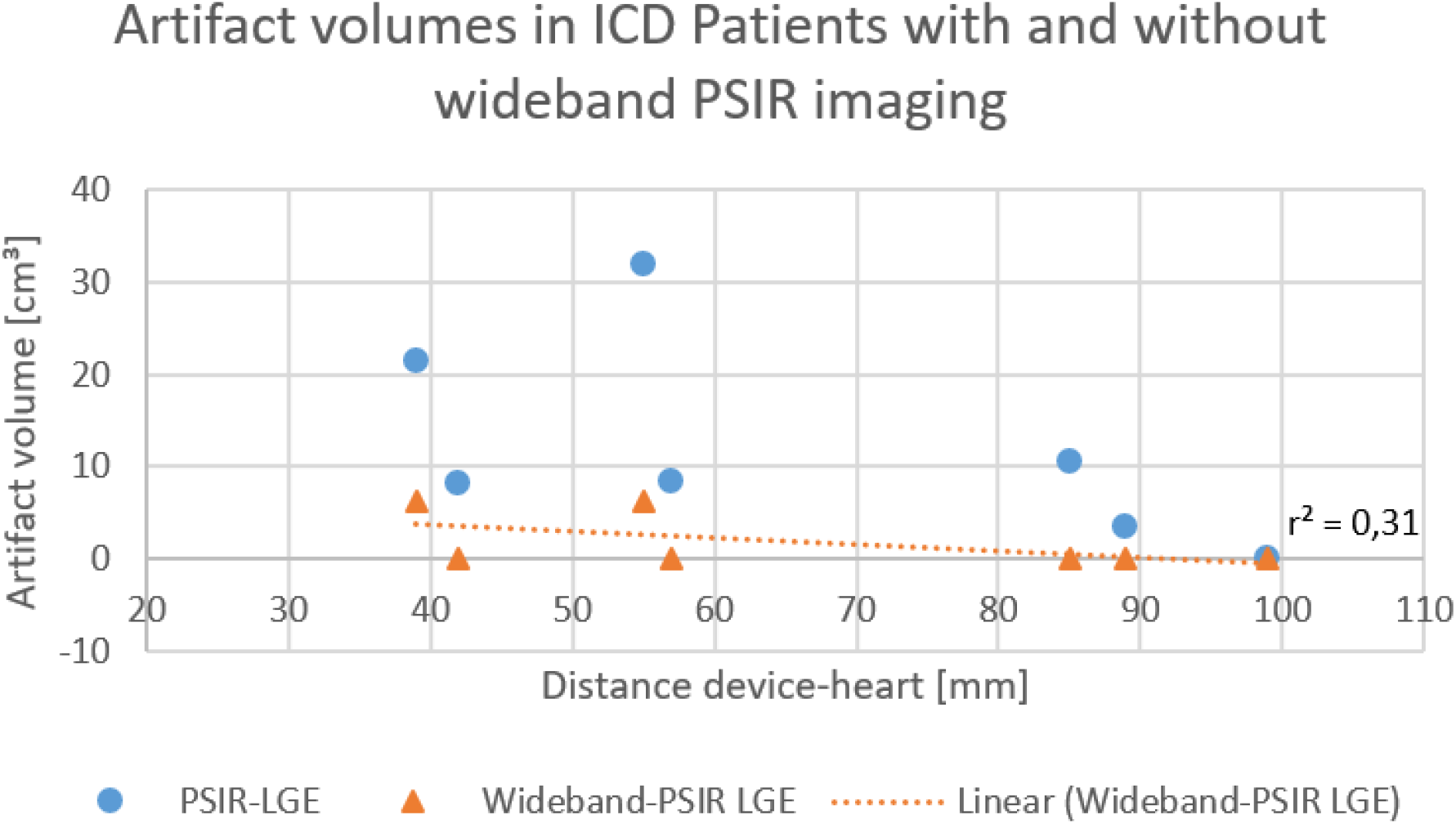
Comparison of artifact burden for wideband PSIR and conventional PSIR imaging. The artifact volume is the sum of the artifact volumes measured in each slice of the full short axis stack. The wideband PSIR imaging successfully suppressed the device’s artifact even in short distance to the heart.

In two patients, the implant’s signal void showed an overlap with the left ventricular anterior wall (distance cardiac shadow - implant 2.9 cm and 5.1 cm, respectively; distance RV lead - implant 12.2 cm and 13.8 cm, respectively) (Fig. 7).

**Fig 7:**
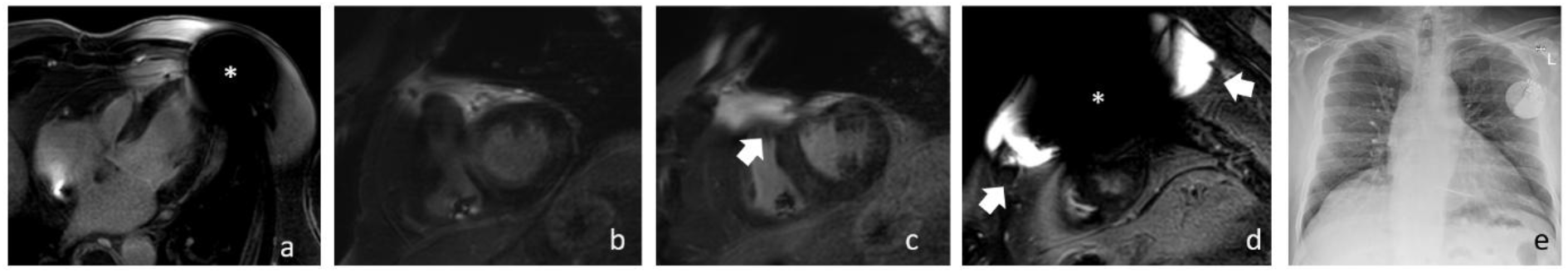
Wideband LGE PSIR imaging with a bandwidth of 3kHz and an offset of +750 Hz in a patient with a 1-chamber ICD (distance from device to heart silhouette 5,4 cm). The signal void (asterisk) reaches the left-ventricular apical segments (a, 4 chamber view). In the close vicinity of the signal void, the wideband LGE PSIR fails to suppress the hyperintense artifact (arrow) while in the other segments the artifact suppression is sufficient for image interpretation (b) basal SA slice, c) midventricular SA slice, d) apical SA slice). e) chest x-ray.

## Discussion

This work shows the feasibility of artifact-reduced CMR imaging of patients with implanted ICD at 3.0T by transferring optimized sequence protocols for CMR from 1.5T.

Adopted from the approach at 1.5T, the use of spoiled gradient echo sequences after administration of gadolinium-based contrast agents removes the majority of ripple artifacts and allows a reliable delineation of endomyocardial structures.(4, 10) Likewise, the application of a wideband inversion pulse for LGE imaging has been shown to counteract the device induced frequency offset that, at 1.5 T easily exceeds the conventional bandwidth of 1 kHz and in general, depends on the device type itself. A scout like approach was proposed, individually determining optimal bandwidth and frequency offset. (12, 14) At 3.0T, both presented phantom measurements and volunteer measurements suggest using a setting with a bandwidth of 3kHz and frequency offset of +750 Hz. The scans of patients with implanted devices highlight the practicability of this approach, widely minimizing artifacts without additional scouts for optimal sequence settings.

As has been shown previously, despite the clear benefit of modified protocols, both for CINE imaging and for LGE imaging, there are remarkable differences in the artifact burden between different devices. (7, 8, 10, 12, 15) A correlation to the distance between the device and the cardiac region has been demonstrated and as an additional countermeasure against artifact overlap, a modified patient positioning with the arm on the side of the device elevated above the head has been proposed. (12, 18, 20) However, considering the wide age range up to 85 years old and constitutional limitations, we omitted any arm rising maneuvers to increase the distance between device and the region of interest that have been discussed as additional tool for an improved image qualityIt has been descripted at 1.5T, and due to the anatomical setup, at 3.0T, the anterior wall remains the most prone to residual artifact burden. (10, 12)

. In recent works at 3.0T, even while using the arm rising approach, artifact burden during CINE imaging was not completely suppressed, and not in all cases this approach was feasible or sufficient to increase the distance adequately. (15, 21) Similar to our findings, the case series of Black and al. revealed a variable artifact burden in comparable distances between device and heart, despite the option of rising the arm during the scans. (15)

In previous phantom works, we demonstrated the different qualities of device induced artifacts, divided by signal void and distortion, the latter characterized by abnormalities in signal intensities, minimizing and maximizing effects as well as out-of round effects. The phantom measurements also revealed not only a dependency of the device type, such as pacemaker or ICD, but also on the inner setup of the device. Especially the signal void does not have a compact spherical appearance but can be divided into smaller spherical structures that are not symmetrical in all spatial directions.(7, 8) This difficult to predict area of artifact burden most likely contributes to the weak correlation of artifact suppression and distance between the device and artifact.

(7, 8)

One of the remaining challenges is the use of vector ECGs at 3.0T in the presence of active implants. According to current recommendations for patient preparations prior to MRI or CMR scans, patients with ICD and recurrent ventricular arrhythmias are considered at high risk and require close monitoring.(22) However, the vector ECG recommended for supervision of patients with active cardiac implants showed a high artifact burden and in fact was not usable neither for this purpose nor for cardiac triggering. From the early days of CMR imaging it is known that both static and varying magnetic fields induce effects such as magneto-hydro-dynamic effects, and in clinical routine countermeasures such as noise cancelling and filter have been introduced. (23) However, the presence of active cardiac implant disturb the existing magnetic fields, most likely inducing additional magento-hydro-dynamic effects, that are not intercepted by the established systems. In this context, a strict limitation of examination time seems reasonable.

## Conclusion

The presented work shows the feasibility of GRE-CINE and wideband PSIR-LGE for cardiac imaging of patients with ICDs at 3.0T Application of contrast agent and GRE CINE imaging reduced the artifact burden from up to 100% to 66% or less. The application of wideband PSIR LGE imaging allowed a complete diagnostic evaluation of myocardial scar in 71% of cases, compared to 14% with conventional LGE imaging without further maneuvers such as arm rising.

## Data Availability

All data produced in the present work are contained in the manuscript

## Limitation

The presented data are limited by the overall small size of the pilot series and the limited number of tested devices.

## Acknowledgement

We would like to thank I. Perdijk for her support in performing the experiments

## Author contributions

T.R. wrote the main manuscript and substantially contributed to the study design and data acquisition. I.W. and O.W. contributed substantially to the study design and data analysis, as well as manuscript revision. N.K., M.H. and S.K. substantially contributed to data acquisition and manuscript revision. W.B. substantially contributed to study design, and manuscript revision. All authors read and approved the final manuscript.

## Data availability

The datasets used and analyzed during the current study are available from the corresponding author on reasonable request (contact).

## Competing interests

Dr. Weiss is a former employee of BIOTRONIK SE & Co. KG, Berlin, Germany, and Dr. Weber is an employee of Philips GmbH, Hamburg, Germany. Prof. Bauer is a scientific advisor for BIOTRONIK SE & Co. KG, Berlin, Germany. Also, this work has been partially funded by BIOTRONIK SE & Co. KG, Berlin, Germany. Dr. Reiter, Dr. Kraus, Dr. Walther and Dr. Kress have no competing interests to declare.

---

**Supplementary Figure 1:**
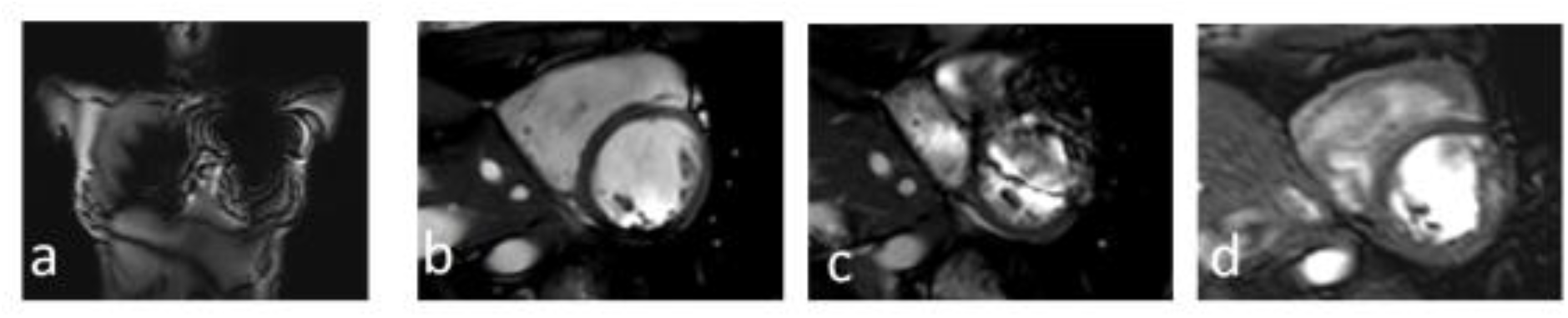
native CINE imaging in a volunteer. a) coronar survey scan with the device taped on the chest. b) bSSFP CINE short axis view without taped on device. c) identical sequence settings as in b) but with taped on device. d) CINE short axis view, TFE-CINE, no flow compensation, partial echo. The contours do not appear as sharply distinguished as with the bSSFP CINE, however, there is an unobstructed view on all wall segments.

**Supplementary Figure 2:**
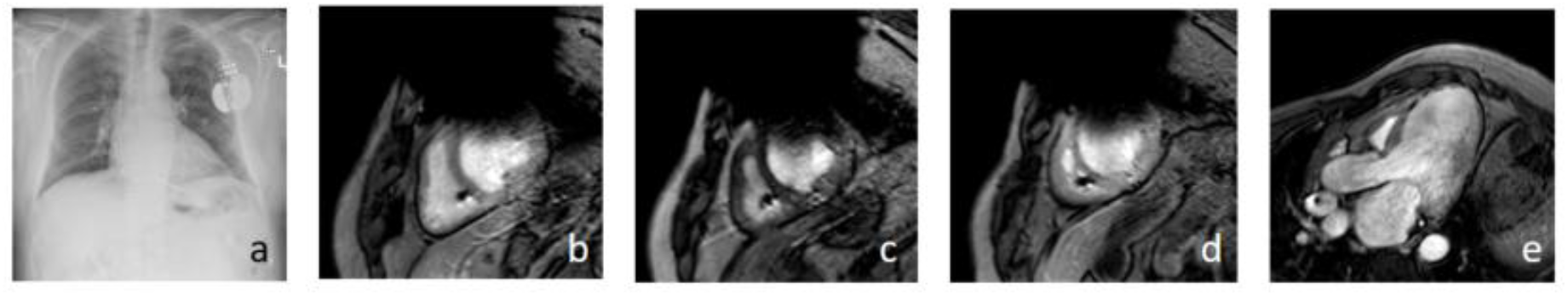
a) shows the short distance between device and the cardiac silhouette. Despite optimized protocol, the CINE SA images (b-d) show significant overlap of the signal void with the anterior wall. Notably, due to the different angulation the 3-Chamber view avoids the signal void completely and shows no aftifact at all.

**Supplementary Figure 3:**
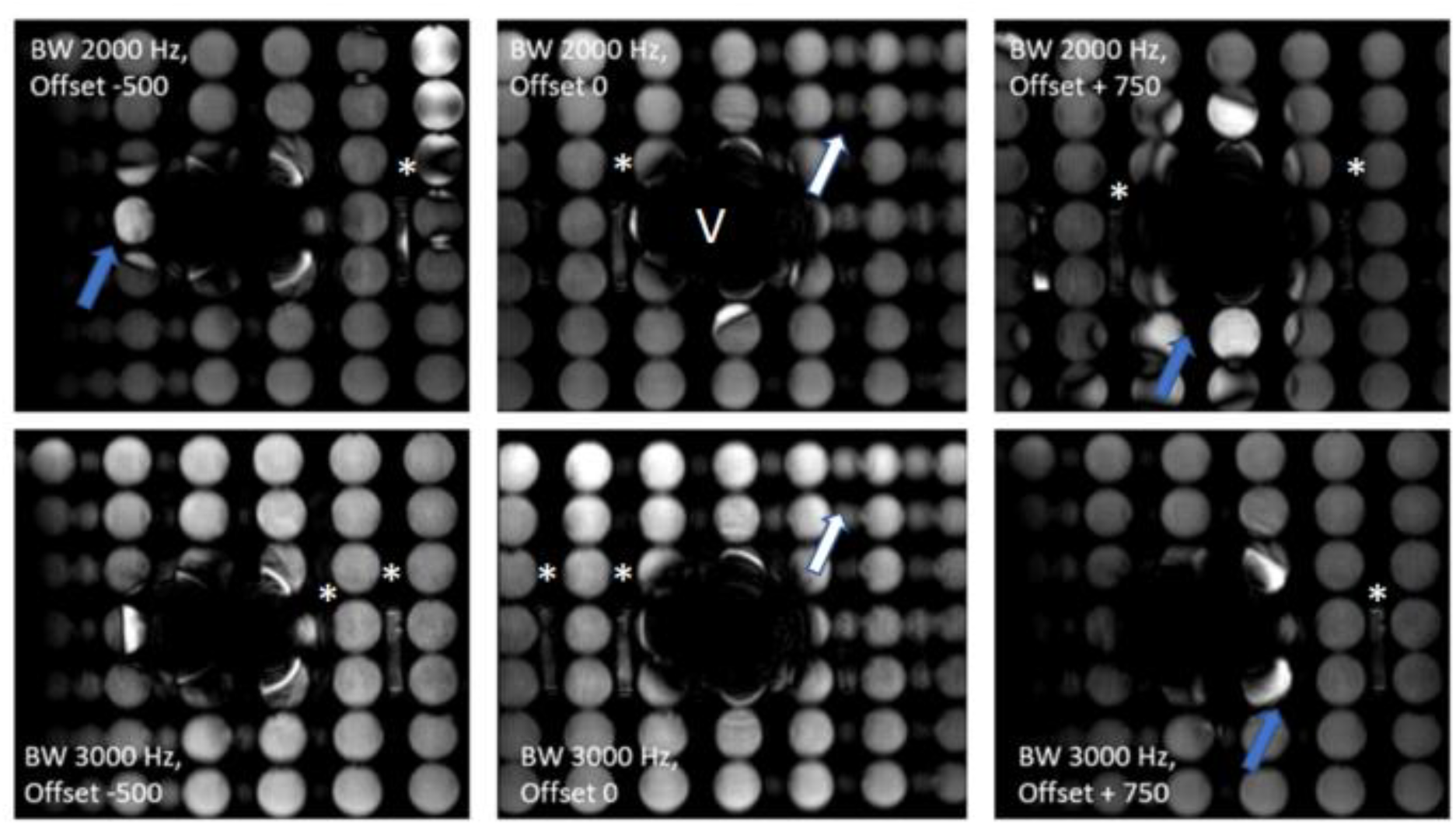
Phantom measurements using the Wideband-PSIR-LGE with adiabatic inversion pulses in the presense of a Biotronik Acticor 7 HF-T. The signal void caused by the device is marked with a with V. Sequence Settings: TR 3ms, TE 1 ms, flip angle 15°, echo train length 49, bandwidths: 2kHz and 3kHz, frequency offsets-500 Hz, 0 and +750 Hz. White asterisks mark the small tubes with contrast agent containing fluid. Blue arrows highlight the areas of hyperintens artifacts. With changes of frequency offset, the position of the hyperintense artifact shifts, eg. from lateral (BW 2kHz, Offset -500 Hz) to inferior (BW 2kHz, Offset + 750Hz). The white arrows mark geomtrical distortions, suggesting additional structures, that acutally are not present. The artifact burden is less at a bandwidth of 3kHz than at 2kHz.

**Supplementary Figure 4:**
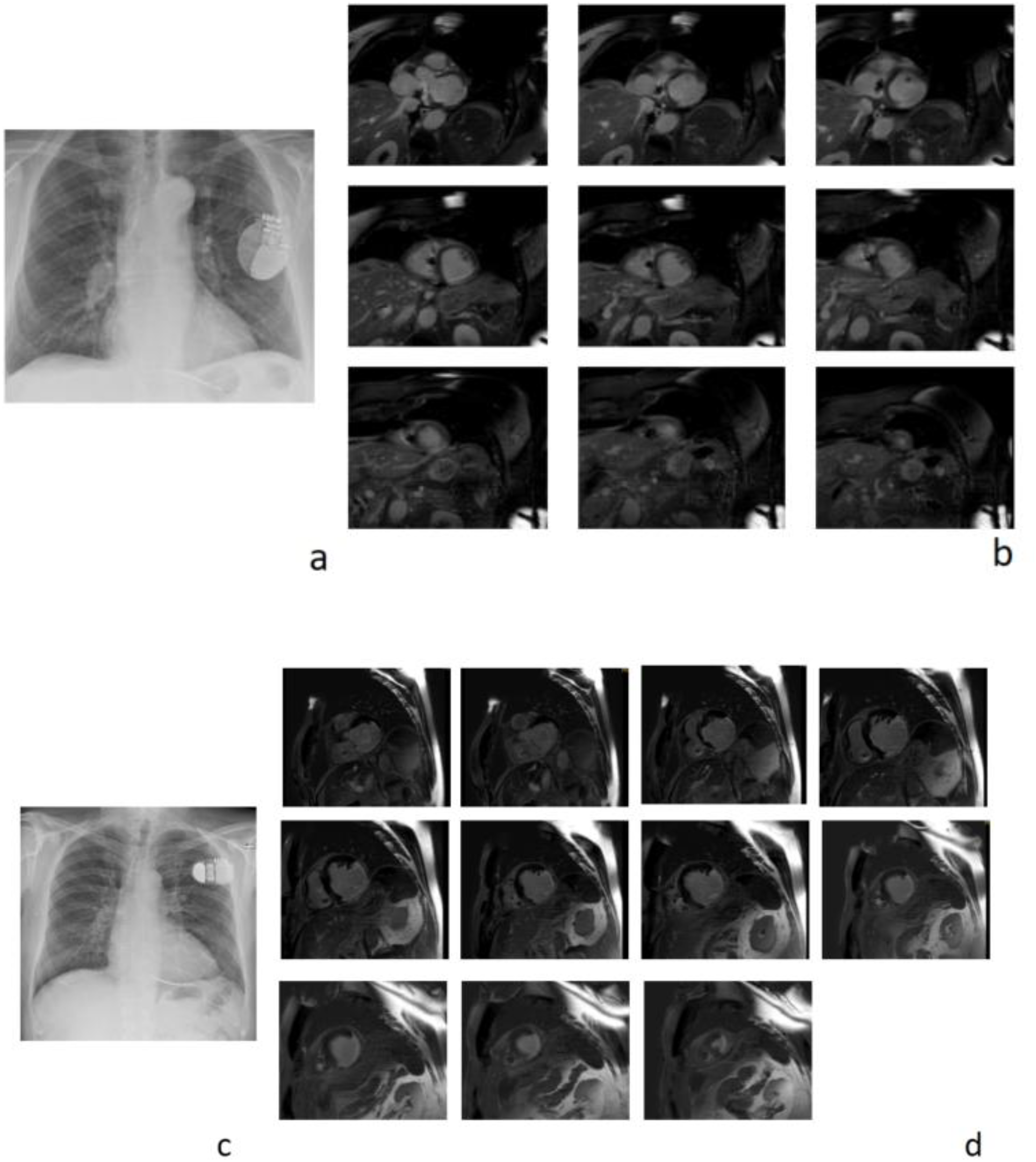
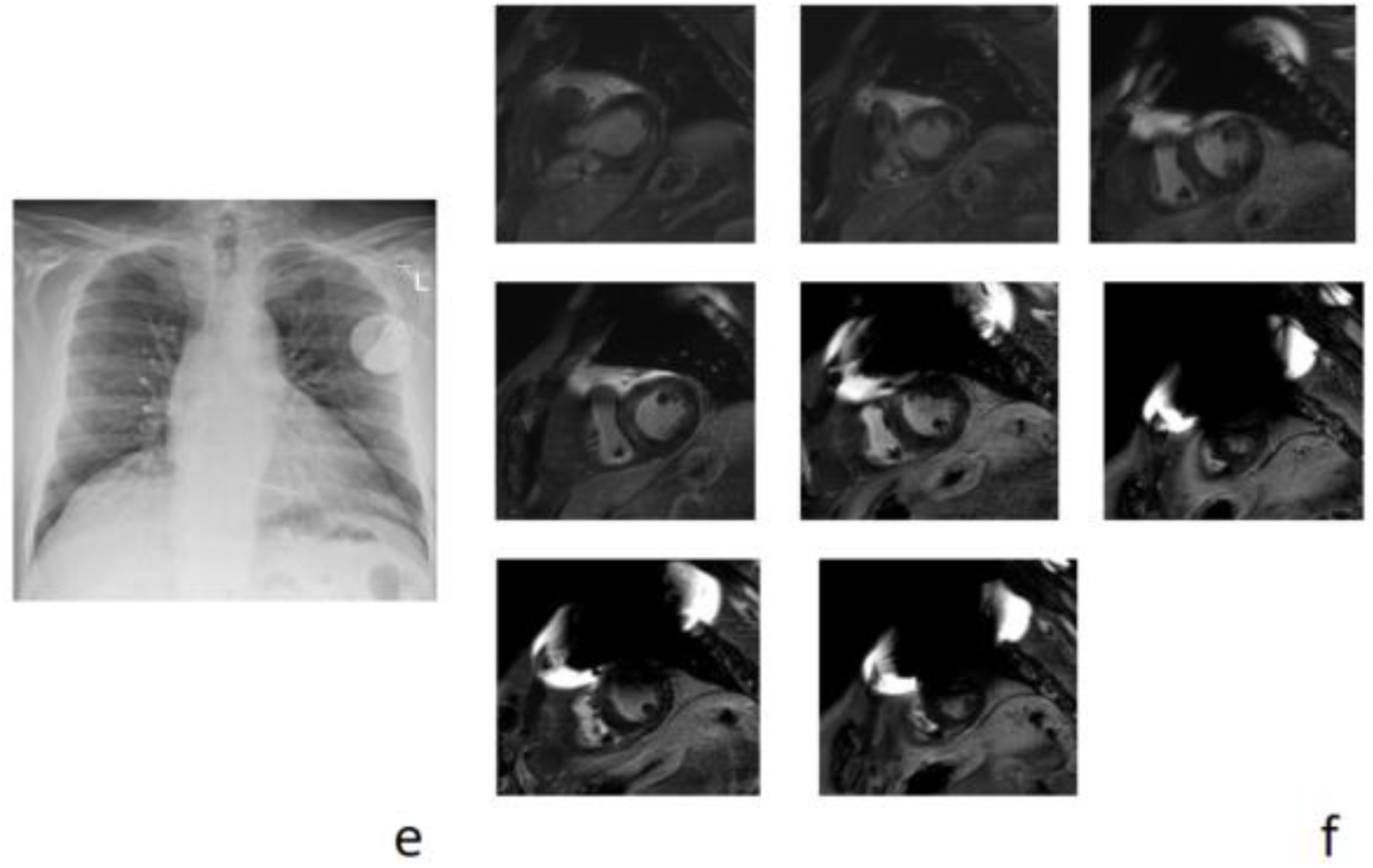
Three exemplary full stacks of Wideband LGE PSIR imaging with a bandwidth of 3kHz and an offset of +750 Hz. Case 1 (a) and b)): patient (range 70-75 years) with ischemic heart disease. The distance between device and cardiac silhouette was 40 mm (a). Case 2 (c) and d)): patient, (range 70-75 years) also in the seventies, with ischemic heart disease, The distance between device and cardiac silhouette was 78 mm (c). Case 3 (e ) and ff)): patient (65-70 years ) with DCM. The distance between device and cardiac silhouette was 54 mm (f).

## Notes

### Author Declarations

The "Medizinische Ethikkommission an der Julius-Maximilians-Universitat Wurzburg" gave full approval for this study

